# Impact of a pilot community pharmacy system redesign on reducing over-the-counter medication misuse in older adults

**DOI:** 10.1101/2019.12.19.19014886

**Authors:** Aaron M. Gilson, Jamie A. Stone, Ashley O. Morris, Roger L. Brown, Ka Z Xiong, Nora Jacobson, Richard J. Holden, Steven M. Albert, Cynthia H. Phelan, Denise L. Walbrandt Pigarelli, Robert M. Breslow, Lauren Welch, Michelle A. Chui

## Abstract

**Objectives:** This pilot study examines effectiveness of an innovative pharmacy design change on over-the-counter (OTC) medication misuse in older adults (ages ≥65). Few interventions have attempted to decrease older adult OTC misuse, and none have addressed system barriers. A structural redesign of the pharmacy (the Senior Section™) was conceptualized to increase awareness of higher-risk OTC medications. The Senior Section contains a curated selection of OTC medications (for pain, cough/cold, allergy, sleep) and is close to the prescription department to facilitate pharmacy staff/patient engagement to reduce misuse.

**Methods:** A pre-/post-implementation design was used to recruit 87 older adults from three pharmacies. Using a hypothetical scenario, participants selected an OTC medication, which was compared to their medication list and health conditions, and their reported use was compared against the product labeling. Four misuse outcomes were determined: (1) Drug/Drug, (2) Drug/Disease, (3) Drug/Age, and (4) Drug/Label with five sub-types. Patient characteristics were collected and compiled into a propensity-score matching logistic regression model to estimate their effects on the Senior Section’s association with misuse outcomes at pre-/post-implementation.

**Results:** Patient characteristic were uniform between pre-/post-implementation and, once entered into a propensity-score matching model, Drug/Disease Misuse significantly lessened over time (z=-2.09, p=0.037). The Senior Section reduced Drug/Drug Misuse, but not significantly. Drug/Label Misuse varied according to the sub-type, with reduced Daily-Dosage (z=-2.42, p=0.016) and Single-Dosage misuse (z=-5.82, p=0.001); however, Timing/Frequency misuse increased (z=2.16, p=0.031).

**Conclusions:** These nascent outcomes support a well-conceived pharmacy-based OTC aisles redesign as valuable for reducing older adult OTC medication misuse. The Senior Section, when broadly implemented, would create new permanent structures and processes to assist older adults in accessing risk information for confidently selecting safer OTC medications.

## Introduction

Over the last 15 years, there has been increasing interest in investigating and mitigating the prevalent problem of misuse of over-the-counter (OTC) medications.^1-7^ The accumulating body of empirical evidence gained from directly investigating OTC misuse coincided with published literature estimating the extent of adverse events among older adults while examining the general incidence of OTC use (see ^8-9^ for example). Assessments of national data indicate that older adults who report that they are currently taking OTC medications are susceptible to using those medications unsafely. For example, while over a third of older adults take at least one OTC medication, more than two-thirds of those concurrently use OTC medications with prescription medications or dietary supplements, which creates a potential for adverse medication interactions.^10^ In addition, OTC-related adverse drug events continue to be implicated in emergency hospitalizations involving older adults.^9^ In many cases, these undesirable health consequences represent an unanticipated consequence of patients’ polypharmacy.^11^

Harms resulting from OTC-related adverse events are often a function of medication misuse, which is uniquely high for the older adult population due to a variety of factors – including increased risks related to certain medications based on a person’s age (representing Drug/Age misuse), interactions with concurrent medications (representing Drug/Drug misuse), exacerbation of current health conditions (representing Drug/Disease misuse), and deviations from recommended usage instructions (representing Drug/Label misuse).^7,12^ Although these situations are possible to identify and address within a primary care setting, practitioners often remain unaware of their patients’ OTC medication use.^1,13^ To further complicate matters, patients frequently lack knowledge about the safety issues associated with the OTC medications that they choose to take without guidance from a healthcare professional,^14-16^ although recent evidence relating to patients in the U.S. is scarce.

Despite the decades-long availability of published resources such as the Beers Criteria,^17-22^ prevention of misuse within the older adult population can be undermined by the types of both health professional and patient issues discussed above. This evidence suggest that practice-based interventions can be an important method in which to reduce the potential for patient harms through the inappropriate use of OTC medications. Unfortunately, clinical practice and research has not traditionally generated testable practice design interventions to address system barriers for decreasing misuse of high-risk OTC medications in older adults.

### The Senior Section ™

To meet this need, the frameworks of participatory design^23^ and human factors engineering^24^ were used to redesign a structural layout of the pharmacy (as described in the study protocol^12^). Community pharmacies were considered an ideal system in which to implement an intervention in an attempt to prevent misuse, given their prominence in the community as an easily-accessible source of OTC medications coupled with pharmacists’ professional training and experience in medication safety. The re-designed structural layout (called the Senior Section) aimed to increase awareness of higher-risk OTC medications, and to promote interactions between pharmacy staff and older adults that would promote safer OTC medication decisions. The Senior Section contains a specialized, curated, layout for OTC medication categories (for pain, cough/cold, allergy, sleep) with lower risk profiles, displays signage showing medication safety warnings, and is close to the prescription department to facilitate pharmacy staff/patient engagement. A fundamental hypothesis of this intervention is that older adult patients who gain insights into potential OTC medication safety issues, either through interactions with pharmacy staff or by following cautionary signage content, will be less likely to misuse OTC medications and thus be at reduced risk of use-related harms.

### Objective

This pilot study was designed to examine the effects of an innovative pharmacy design change on the reported misuse of OTC medications by older adults. Two research questions guided this study: (1) Did implementing the Senior Section in a small sample of community pharmacies reduce occurrence of Drug/Age, Drug/Drug, Drug/Disease, and Drug/Label misuse, and (2) Did various patient characteristics influence the Senior Section’s effect on these misuse outcomes? It was hypothesized that innovation implementation would decrease the frequency of OTC medication misuse, and that patient characteristics had the potential to moderate this effect.

## Methods

A pre-/post-implementation design was used to assess changes in the occurrence of misuse for older adults recruited from a small sample of community pharmacies within a single pharmacy organization.

### Recruitment

Three community pharmacies from the same pharmacy chain were selected for project participation. Each pharmacy location generated a list of customers who were 65 years or older and received a prescription from that pharmacy within the last year. Older adults received an invitation letter to participate in the study, sent from the pharmacy manager at each site, explaining that the store was collaborating with University of Wisconsin researchers to learn more about how older adults select and use OTC medications. The letter stated that eligible participants were within a certain age category and had either purchased or considered purchasing an OTC medication in the past year to treat either pain, insomnia/sleep problem, cough/cold, or allergies. A decision was made to include all community-dwelling older adults, regardless of their cognitive capacity. Although a range of cognitive capacity could be expected, exclusions were not made because all non-institutionalized individuals comprise the OTC medication-purchasing population.

A cumulative total of 1350 letters were mailed to eligible participants during the pre-implementation phase, while 450 were mailed to different participants during the post-implementation phase. Study fliers also were placed near the pick-up window at each pharmacy. Both letters and fliers contained instructions for potential participants to contact the study team to learn more about the study. Pharmacy staff could also call the study team with contact information for potential participants, if that older adult preferred to be contacted directly. Interested participants were then phoned to be given information about the study requirements and, if desired, a date and time was scheduled to complete an interview at the pharmacy location at which they were a regular customer. Study materials were mailed prior to the interview. Participants were paid $20 for completing the study. This recruitment method resulted in the selection of two separate cohorts – one for pre-implementation and one for post-implementation. Recruitment occurred from March to September 2018. This study is a component of a larger research project that was approved by the University of Wisconsin Institutional Review Board.

Post-implementation recruitment and patient participation was severely hampered by the unanticipated and rapid closure of the pharmacy chain, which occurred during the project timeframe. Although an effort was made to accelerate recruitment efforts after the closure was announced, there was insufficient time to accumulate the anticipated number of completed patient interviews. As a result, the post-implementation sample size was lower than originally predicted, resulting in notably unequal sample sizes between time periods. The Statistical Analysis section below outlines the approach taken to ensure appropriate statistical comparisons of these disparate sample sizes.

### Data Collection

Participants completed one pharmacy-based interview and two questionnaires. Prior to the interview, participants were mailed a questionnaire that assessed nine patient characteristics – health status (Likert scale, 1=poor, 5=excellent), health conditions (the Older Americans Resource Survey methodology), 30-day medication use (self-reported for prescriptions, OTCs, and dietary/herbal supplements), number of prescribers and pharmacies (self-reported), age, gender, education, and race. Participants brought the completed questionnaire with them to their scheduled interview. All participants (either pre-or post-implementation) completed the same data collection tools.

Participants were met by the interviewer near the store entrance. After collecting study materials and answering any participant questions, the participant was asked to choose one of the following symptom scenarios that applied to them:

Scenario 1: Sleep

> *Recently, you have been having (more) difficulty falling asleep or staying asleep. You are here at [name of store] to look for a medication that can help you sleep*.

Scenario 2: Pain

> *You are having a soreness and muscle aches. It is not bad enough to call your doctor. You have not taken any medication to help with these aches yet. You are here at [name of store] to look for a medication that can help you feel better*.

Scenario 3: Cough/Cold or Allergy

> *For the past three days you’ve had a runny nose, sore throat, felt “stuffy”, and your head is congested. You don’t have a fever and it is not bad enough to call your doctor. You have not taken any medication for your symptoms yet, but you are here at [name of store] to look for a medication that can help you feel better*.

Participants were then asked to show and tell the researcher how they would address the health issue described in the scenario by walking the interviewer through the store to the medication they would select. Occasional probes were offered during the interview, which included questions such as “How did you decide to pick this particular medication?” or “What are you thinking about when you decided to pick this medication instead of other ones?” After selecting a medication, participants were asked to describe how they would use the medication they selected, including such questions as “How would you take this medication? How often? And what time of day?” A short satisfaction survey, assessing patients’ interactions with pharmacy staff and about their OTC medication selection, was administered following the interview. Interviews were audio and video recorded and transcribed. On average, the in-person interview took about 30 minutes to complete. All pre- and post-implementation data were collected between July and December, 2018.

### Misuse Analysis

Three pharmacists with clinical experience working with the geriatric population comprised a misuse analysis team. Prior to the misuse evaluation, information about the participant and OTC medication selected was de-identified and entered into Redcap. This information included participant’s self-reported medication list and health conditions, and the selected OTC and reported OTC use (which included direct portions of the interview transcript and use summaries prepared by the research team). Data were extracted from the transcripts in this manner to ensure that the misuse analysis team would be unable to identify whether the interview occurred before or after Senior Section implementation. Also included were photographs of the OTC medication (front, back, and top to capture all product labeling information) as reference for the misuse analysis team. A random number generator was applied to the participant list, which randomly assigned all participants to three different batches for evaluation. The overall process of randomization and blinding ensured that the misuse analysis team would be unable to identify whether the interview occurred pre- or post-implementation. Misuse was first evaluated independently by each reviewer, and responses were then consolidated in preparation for group discussion and discrepancies between reviewers were noted. The misuse study team, supported by study researchers (JAS, MAC, and KZX) then met as a group to review independent comments, facilitate discussion about discrepancies (which were commonly the result of misuse analysis team members’ interpretation of the drug facts information on the label or what patients meant when describing their use), and achieve consensus about final misuse classifications.

Four misuse outcomes were operationalized by the misuse analysis team through these group discussions:

1. <u>Drug-Age Misuse</u> was identified using the 2015 Beers Criteria list for older adults, where any selected OTC medication included on the list is considered misuse, except NSAIDs that are only used to temporarily treat acute pain.
2. <u>Drug-Drug Misuse</u> was measured using LexiComp risk ratings of medication interactions, and resulted in the following domains that carried enough risk to be considered misuse:
  a. Type C (monitor therapy),
  b. Type D (consider therapy modification), and
  c. Type X (avoid combination).
3. <u>Drug-Disease Misuse</u> was determined by identifying potential interactions between medications and disease states designated as high-risk in Beers Criteria, a condition listed in product labeling, or other (e.g. clinical knowledge of the pharmacist).
4. <u>Drug-Label Misuse</u> considers the following deviations from the product labeling recommendations:
  a. over daily dosage,
  b. exceeds single dose,
  c. dose timing/frequency,
  d. use duration, and
  e. inappropriate indication.

For Drug-Age, Drug-Disease, and Drug-Label misuse, the final determinations were based on evaluations and agreement by the misuse analysis team, and was measured as the frequency of misuse per participant.

### Statistical Analysis

Previous sample size power computations estimated a total of 144 different participants (72 older adults at both pre- and post-implementation) were needed to determine changes in OTC medication misuse.^12^ However, effect size calculations using the current sample sizes (see Results section) revealed at least medium effect sizes^25^ for 7 of the 11 misuse comparisons evaluated at pre- and post-implementation for this study. This metric suggests that the current overall sample size for this study is sufficient to warrant the planned use of test statistics.

To estimate binary treatment effects in a non-experimental statistical setting, when units’ non-random assignment to treatment is due to selection based on observations, reweighting is a valuable approach.^26^ That is, when the treatment is not randomly assigned, it is expected that the treated and untreated units present very different distributions of their observable characteristics. To account for this assumption, an initial propensity score was estimated based on the treatment condition using a Logit model to compute the predicated probability (*π*). Using the pi score, the following weights were constructed: 1/*π* for the treated observations, and 1/(1-*π*) for the untreated observations. It was then possible to calculate the average treatment effect by comparing the weighted means of the two groups. All estimates were conducted using the “teffects” and “treatrew” routines in Stata v.16.^27^ Logistic regression initially was used to determine similarity of patient characteristics at pre/post-implementation of the Senior Section. These variables were then compiled into a propensity-score matching model to estimate their combined effects on the Senior Section’s association with various misuse types.

## Results

Findings are based on recruitment rates of 5% at pre-implementation (72/1350) and 3% at post-implementation (15/450), although the post-implementation rate was influenced by store closures. The status of patient demographic characteristics is provided in Table 1, and revealed few notable differences in patient samples between pre- and post-implementation. At pre-implementation, no patient had an educational level below high school, but one patient reported education up to eighth grade at post-implementation. The number of medications that patients reported taking also was narrower at post-implementation (pre-implementation min/max: 1-33 vs. post-implementation min/max: 6-18), but this did not translate into any distinction between means. In addition, over 60% of patients had a total health rating of very good or excellent prior to Senior Section implementation, while only a third of patients had the same rating at post-implementation. Despite these slight differences, Table 2 shows that, when examined individually, no patient characteristic varied significantly between pre-/post-implementation. Such findings suggest that the patient samples can be considered homogeneous for these characteristics across the two assessment end-points.

**Table 1:**
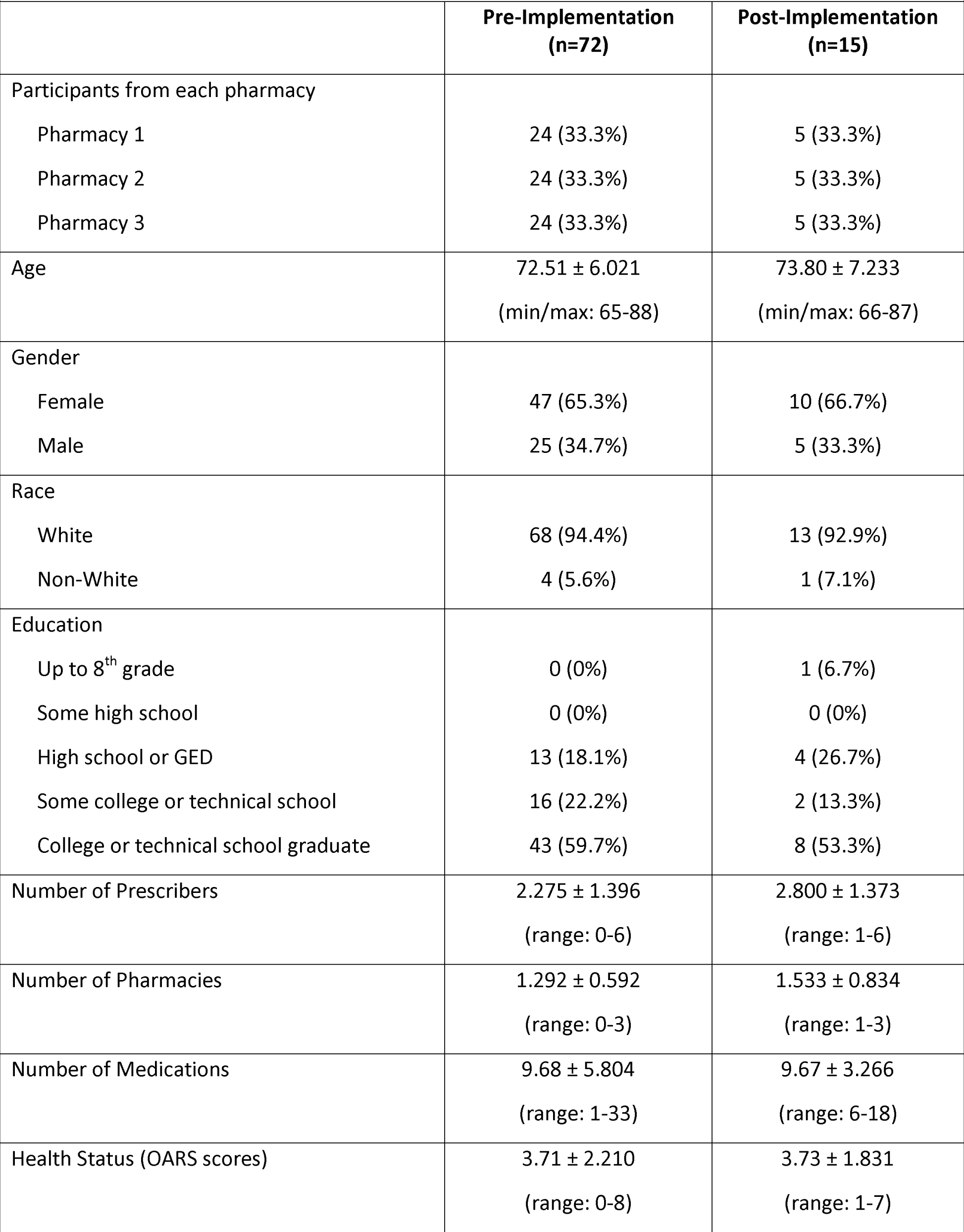
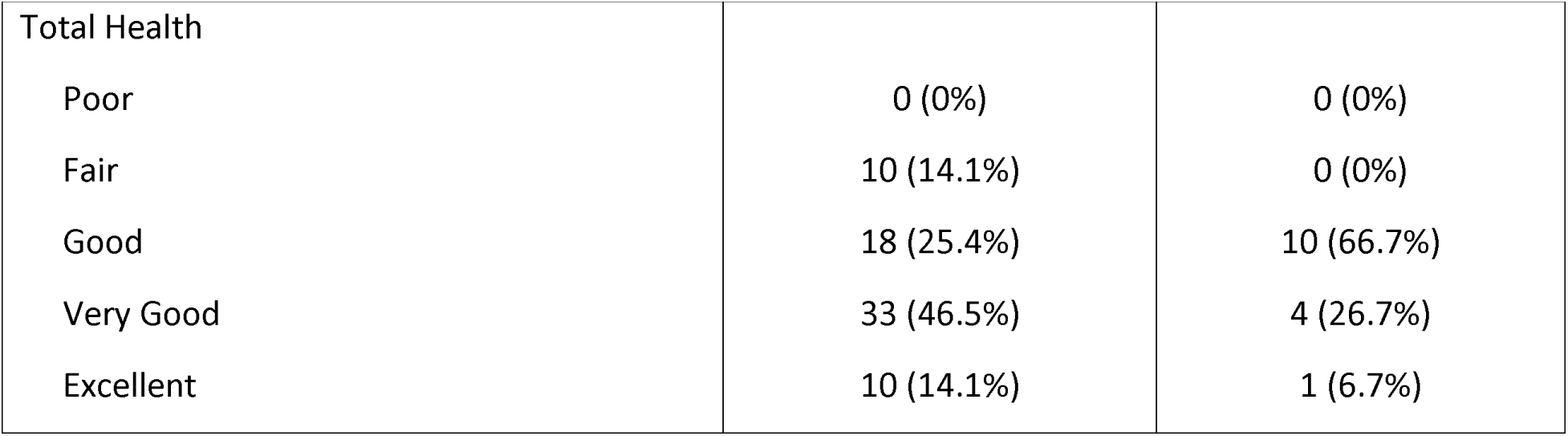
Patient Demographic Characteristics (Pre-Implementation and Post-Implementation)

|  |  |  |
| --- | --- | --- |
|  | **Pre-Implementation**<br>**(n=72)** | **Post-Implementation**<br>**(n=15)** |
| Participants from each pharmacy |  |  |
| Pharmacy 1 | 24 (33.3%) | 5 (33.3%) |
| Pharmacy 2 | 24 (33.3%) | 5 (33.3%) |
| Pharmacy 3 | 24 (33.3%) | 5 (33.3%) |
| Age | 72.51 ± 6.021<br>(min/max: 65-88) | 73.80 ± 7.233<br>(min/max: 66-87) |
| Gender |  |  |
| Female | 47 (65.3%) | 10 (66.7%) |
| Male | 25 (34.7%) | 5 (33.3%) |
| Race |  |  |
| White | 68 (94.4%) | 13 (92.9%) |
| Non-White | 4 (5.6%) | 1 (7.1%) |
| Education |  |  |
| Up to 8<sup>th</sup> grade | 0 (0%) | 1 (6.7%) |
| Some high school | 0 (0%) | 0 (0%) |
| High school or GED | 13 (18.1%) | 4 (26.7%) |
| Some college or technical school | 16 (22.2%) | 2 (13.3%) |
| College or technical school graduate | 43 (59.7%) | 8 (53.3%) |
| Number of Prescribers | 2.275 ± 1.396<br>(range: 0-6) | 2.800 ± 1.373<br>(range: 1-6) |
| Number of Pharmacies | 1.292 ± 0.592<br>(range: 0-3) | 1.533 ± 0.834<br>(range: 1-3) |
| Number of Medications | 9.68 ± 5.804<br>(range: 1-33) | 9.67 ± 3.266<br>(range: 6-18) |
| Health Status (OARS scores) | 3.71 ± 2.210<br>(range: 0-8) | 3.73 ± 1.831<br>(range: 1-7) |
| Total Health |  |  |
| Poor | 0 (0%) | 0 (0%) |
| Fair | 10 (14.1%) | 0 (0%) |
| Good | 18 (25.4%) | 10 (66.7%) |
| Very Good | 33 (46.5%) | 4 (26.7%) |
| Excellent | 10 (14.1%) | 1 (6.7%) |

**Table 2.** Logistic Regression Comparing Patient Characteristics in Pre- and Post-Implementation Samples (n=87)

|  | Odds Ratio | Stand. Error | z | P> \| z \| | 95% Confidence Interval |  |
| --- | --- | --- | --- | --- | --- | --- |
|  |  |  |  |  | Lower Bound | Upper Bound |
| Health Status | 1.021 | 0.441 | 0.05 | 0.962 | 0.437 | 2.382 |
| Total Health | 0.880 | 0.159 | -0.71 | 0.479 | 0.618 | 1.254 |
| Age | 0.990 | 0.053 | -0.19 | 0.846 | 0.892 | 1.098 |
| Gender (male) | 0.715 | 0.510 | -0.47 | 0.639 | 0.177 | 2.897 |
| Education | 0.621 | 0.214 | -1.38 | 0.168 | 0.315 | 1.222 |
| Race (non-white) | 1.119 | 1.420 | 0.09 | 0.929 | 0.093 | 13.463 |
| No. of Prescribers | 1.518 | 0.413 | 1.54 | 0.125 | 0.891 | 2.588 |
| No. of Pharmacies | 1.530 | 0.688 | 0.94 | 0.345 | 0.633 | 3.696 |
| No. of Medications | 0.998 | 0.072 | -0.03 | 0.980 | 0.867 | 1.149 |

Table 3 demonstrates differences between the means for the various types of misuse before the Senior Section was implemented compared to post-implementation. For these samples, the means for OTC medication misuse frequencies tended to be low overall, with only one mean (for Drug/Drug Misuse-Type C at pre-implementation) approaching 1.50 and most being below 0.50. Seven of the 11 comparisons also showed expected patterns of effects, where the frequency of misuse at post-implementation was lower than at pre-implementation. However, for the four types of misuse with higher means after intervention implementation, the difference between means was no more than 0.073.

**Table 3.**
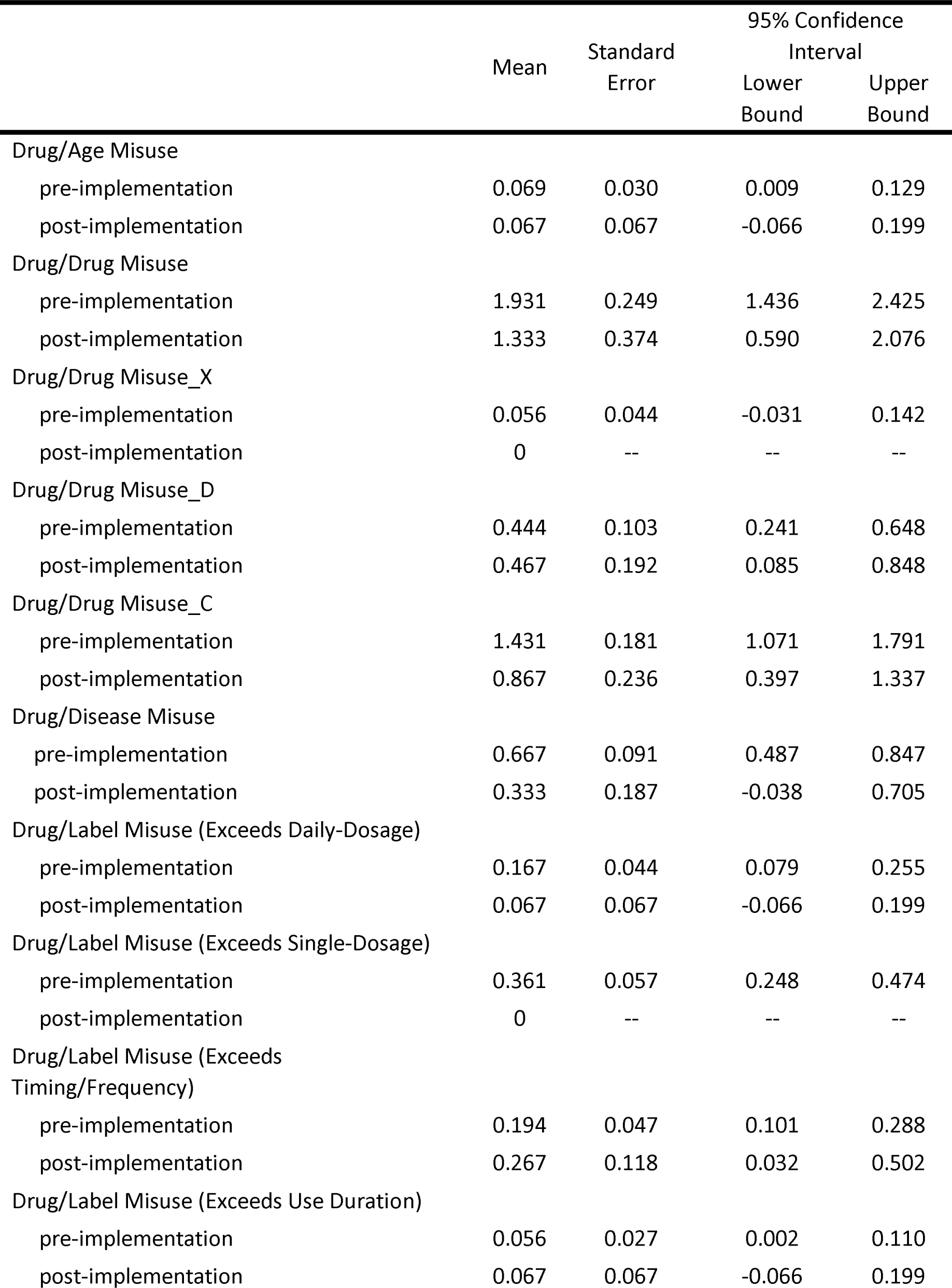
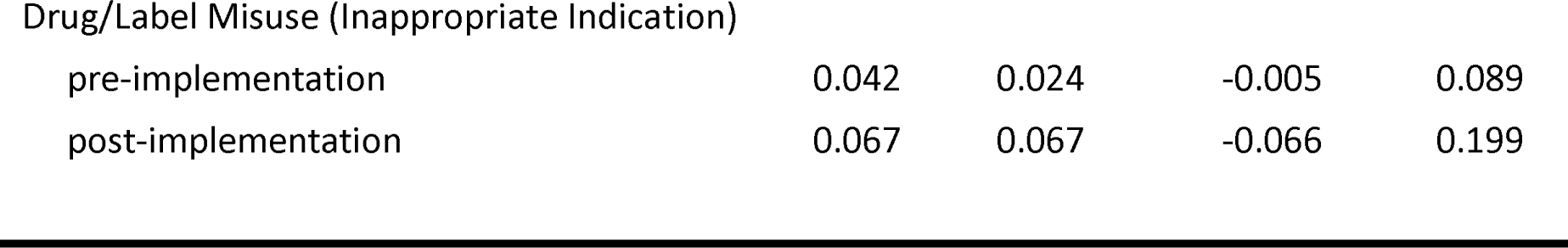
Frequency of Types of Misuse: Pre-Implementation Compared to Post-Implementation (n=87)

|  | Mean | Standard Error | 95% Confidence Interval |  |
| --- | --- | --- | --- | --- |
|  |  |  | Lower Bound | Upper Bound |
| Drug/Age Misuse |  |  |  |  |
| pre-implementation | 0.069 | 0.030 | 0.009 | 0.129 |
| post-implementation | 0.067 | 0.067 | -0.066 | 0.199 |
| Drug/Drug Misuse |  |  |  |  |
| pre-implementation | 1.931 | 0.249 | 1.436 | 2.425 |
| post-implementation | 1.333 | 0.374 | 0.590 | 2.076 |
| Drug/Drug Misuse_X |  |  |  |  |
| pre-implementation | 0.056 | 0.044 | -0.031 | 0.142 |
| post-implementation | 0 | -- | -- | -- |
| Drug/Drug Misuse_D |  |  |  |  |
| pre-implementation | 0.444 | 0.103 | 0.241 | 0.648 |
| post-implementation | 0.467 | 0.192 | 0.085 | 0.848 |
| Drug/Drug Misuse_C |  |  |  |  |
| pre-implementation | 1.431 | 0.181 | 1.071 | 1.791 |
| post-implementation | 0.867 | 0.236 | 0.397 | 1.337 |
| Drug/Disease Misuse |  |  |  |  |
| pre-implementation | 0.667 | 0.091 | 0.487 | 0.847 |
| post-implementation | 0.333 | 0.187 | -0.038 | 0.705 |
| Drug/Label Misuse (Exceeds Daily-Dosage) |  |  |  |  |
| pre-implementation | 0.167 | 0.044 | 0.079 | 0.255 |
| post-implementation | 0.067 | 0.067 | -0.066 | 0.199 |
| Drug/Label Misuse (Exceeds Single-Dosage) |  |  |  |  |
| pre-implementation | 0.361 | 0.057 | 0.248 | 0.474 |
| post-implementation | 0 | -- | -- | -- |
| Drug/Label Misuse (Exceeds Timing/Frequency) |  |  |  |  |
| pre-implementation | 0.194 | 0.047 | 0.101 | 0.288 |
| post-implementation | 0.267 | 0.118 | 0.032 | 0.502 |
| Drug/Label Misuse (Exceeds Use Duration) |  |  |  |  |
| pre-implementation | 0.056 | 0.027 | 0.002 | 0.110 |
| post-implementation | 0.067 | 0.067 | -0.066 | 0.199 |
| Drug/Label Misuse (Inappropriate Indication) |  |  |  |  |
| pre-implementation | 0.042 | 0.024 | -0.005 | 0.089 |
| post-implementation | 0.067 | 0.067 | -0.066 | 0.199 |

When entering these combined characteristics into a propensity-score matching model, statistical effects also differed as a function of the type of misuse (see Table 4). Regardless of whether the patient characteristics propensity score was entered into the model, the Senior Section intervention did not significantly change either Drug/Age Misuse or the three elements of Drug/Drug Misuse (C, D, and X), although there were fewer instances of most of these behaviors at post-implementation. As such, the cumulative covariates had little if any adjusting effect on those outcomes. Alternatively, instances of Drug/Disease Misuse significantly lessened over time only when considering the influence of the patient characteristics (z=-2.09, p=0.037).

**Table 4.** Examining Types of Misuse Using Regression Models with Propensity-Score Matching (PS): Pre- Implementation Compared to Post-Implementation (n=87)

| Models with or without PS | Coef. | Stand.<br>Error | z | P> \|z\| | 95% Confidence<br>Interval |  |
| --- | --- | --- | --- | --- | --- | --- |
|  |  |  |  |  | Lower<br>Bound | Upper<br>Bound |
| Drug/Age Misuse |  |  |  |  |  |  |
| (post vs. pre) | -0.003 | 0.071 | -0.04 | 0.969 | -0.142 | 0.136 |
| (PS, post vs. pre) | -0.049 | 0.047 | -1.05 | 0.294 | -0.142 | 0.043 |
| Drug/Drug Misuse |  |  |  |  |  |  |
| (post vs. pre) | -0.597 | 0.438 | -1.36 | 0.172 | -1.455 | 0.260 |
| (PS, post vs. pre) | -0.457 | 0.372 | -1.23 | 0.220 | -1.186 | 0.273 |
| Drug/Drug Misuse_X |  |  |  |  |  |  |
| (post vs. pre) | -0.556 | 0.043 | -1.28 | 0.201 | -0.141 | 0.030 |
| (PS, post vs. pre) | -0.049 | 0.043 | -1.14 | 0.255 | -0.134 | 0.036 |
| Drug/Drug Misuse_D |  |  |  |  |  |  |
| (post vs. pre) | 0.022 | 0.212 | 0.11 | 0.916 | -0.392 | 0.437 |
| (PS, post vs. pre) | 0.049 | 0.151 | 0.33 | 0.743 | -0.246 | 0.345 |
| Drug/Drug Misuse_C |  |  |  |  |  |  |
| (post vs. pre) | -0.564 | 0.291 | -1.94 | 0.052 | -1.133 | 0.006 |
| (PS, post vs. pre) | -0.457 | 0.267 | -1.71 | 0.087 | -0.980 | 0.066 |
| Drug/Disease Misuse |  |  |  |  |  |  |
| (post vs. pre) | -0.333 | 0.202 | -1.65 | 0.098 | -0.729 | 0.620 |
| (PS, post vs. pre) | -0.407 | 0.195 | -2.09 | 0.037 | -0.790 | -0.248 |
| Drug/Label Misuse (Exceeds Daily-Dosage) |  |  |  |  |  |  |
| (post vs. pre) | -0.100 | 0.080 | -1.28 | 0.200 | -0.253 | 0.053 |
| (PS, post vs. pre) | -0.160 | 0.066 | -2.42 | 0.016 | -0.291 | -0.030 |
| Drug/Label Misuse (Exceeds Single-Dosage) |  |  |  |  |  |  |
| (post vs. pre) | -0.361 | 0.057 | -6.38 | 0.001 | -0.472 | -0.250 |
| (PS, post vs. pre) | -0.358 | 0.062 | -5.82 | 0.001 | -0.479 | -0.273 |
| Drug/Label Misuse (Exceeds<br>Timing/Frequency) |  |  |  |  |  |  |
| (post vs. pre) | 0.072 | 0.123 | 0.59 | 0.558 | -0.170 | 0.314 |
| (PS, post vs. pre) | 0.284 | 0.132 | 2.16 | 0.031 | 0.026 | 0.542 |
| Drug/Label Misuse (Exceeds Use Duration) |  |  |  |  |  |  |
| (post vs. pre) | 0.011 | 0.070 | 0.16 | 0.874 | -0.126 | 0.148 |
| (PS, post vs. pre) | 0 | 0.061 | 0.00 | 1.000 | -0.120 | 0.120 |
| Drug/Label Misuse (Inappropriate Indication) |  |  |  |  |  |  |
| (post vs. pre) | 0.025 | 0.069 | 0.36 | 0.715 | -0.109 | 0.159 |
| (PS, post vs. pre) | -0.012 | 0.025 | -0.49 | 0.621 | -0.061 | 0.037 |

For Drug/Label Misuse, the Senior Section’s influence seemed to vary according to the sub-type, with Daily-Dosage Misuse achieving significant reductions only with the patient characteristics accounted for in the model (z=-2.42, p=0.016). Single-Dosage Misuse decreased after the intervention, as represented in both regression models (without propensity score: z=-6.38, p=0.001; with propensity score: z=-5.82, p=0.001). However, when the effects of patient characteristics were added as a propensity score, Timing/Frequency Misuse became significantly more frequent at post-implementation (z=2.16, p=0.031). Exceeding medication use duration or using medication for an inappropriate indication were only slightly more frequent at post-implementation, but this pattern of effect changed for Inappropriate Indication Misuse when the propensity score was introduced to the model.

## Discussion

Results suggest that a simple but well-conceived redesign of the OTC aisles in a small number of community pharmacies can reduce some categories of older adult OTC medication misuse. Comparing data from homogenous samples over time, these analyses suggest that the Senior Section influenced the frequency of misuse, with the degree of change depending on the specific type of misuse and patient characteristic effects. The two misuse categories that seemed most responsive to the system redesign were Drug/Disease Misuse and Drug/Label Misuse. Drug/Disease Misuse, Drug/Label Misuse (Exceeds Daily-Dosage), and Drug/Label Misuse (Exceeds Timing/Frequency) became significant only after the patient demographic covariate was added to the models; the covariates comprising the patient demographic propensity score had an adjusting impact, making the effect of the intervention more sensitive. However, the occurrence of Drug/Label Misuse (Exceeds Timing/Frequency) challenged expectations by being reported more frequently after the Senior Section was implemented. Despite this single significant finding, almost 70% of all misuse comparisons (from all models, either excluding and including the propensity score) were trending in the direction of anticipated effects, with lower frequencies occurring at post-implementation.

Although it was originally hypothesized that the Senior Section would have a notable diminishing effect on all types of misuse, in retrospect there are clear reasons that may argue against the validity of this expectation. Drug/Age Misuse, based on the Beers Criteria list, likely did not change statistically because of two factors. First, ibuprofen remained in the Senior Section inventory because of its benefits for treating a variety of symptoms, even though the Beers Criteria contains a recommendation to avoid its chronic use for pain management. Second, it was often difficult to ascertain when this misuse type was occurring, because a determination was frequently needed to be made between a patient’s acute and chronic use. Alternatively, for Drug/Drug Misuse, the cautionary signage contained in the Senior Section did not include warnings about specific drugs or about potentials for interactions. When expanding implementation of the Senior Section into a broader network of pharmacies, which is being planned, modifications will be necessary to determine the best approach to address a greater variety of misuse types.

Overall, these were encouraging patterns of effects, and were demonstrated even given the relatively small post-implementation sample size. This intervention, if more broadly implemented in other pharmacy corporations, would create new permanent structures and processes that could improve the quality and availability of information for older adults as they approach the OTC aisles. Such information could lead to greater risk awareness, and help older adults more easily determine their own risk levels and to select safer OTC medications with confidence. Further research must evaluate the generalizability of the intervention’s results and the sustainability of post-implementation improvements in different pharmacy environments. Physically redesigning OTC aisles may also be tested in different vulnerable populations, such as pediatric patients. Taken together, these preliminary outcomes support the Senior Section as a valuable tool for pharmacy staff to improve patients’ safe OTC medication use through heightened awareness and education efforts.

### Strengths & Limitation

The principal strength of this pilot study was its innovation. It was the first to empirically evaluate implementation of a pharmacy redesign intervention on the prevalence of the reported misuse of OTC medications in homogenous samples of older adults, as well as evaluating OTC selection and use within a naturalistic setting.

Despite the crucial insights gained from this analysis, a various limitations warrant consideration beyond the limited generalizability of this small sample of community pharmacies. First, a number of patient characteristics were self-reported (e.g., health status, 30-day medication use, and number of prescribers and pharmacies seen), so future data collection efforts should attempt to employ existing health systems databases. Second, although the patient sample size was limited (n=87 cumulatively between pre- and post-implementation). However, the statistical approach was chosen specifically to best accommodate this sample size and compute valid results. Third, throughout this study, patients were not randomly selected but rather were chosen through recruitment methods. Fourth, patient interview responses were based on reactions to a hypothetical health scenario and may not represent “real world” behaviors. Fifth, results could vary based on which scenario the participant selected (i.e., pain, cough/cold, allergy, or sleep). Additional research is necessary on a larger selection of participants to determine the influence of medication category on types of misuse. Finally, as mentioned previously, the Senior Section may not be designed sufficiently to address certain types of misuse, such as Drug/Drug Misuse and Drug/Age Misuse, and future research should consider the intervention features constructed specifically to reduce a broader array of misuse.

## Conclusions

This pilot study provides initial insights into the extent that a pharmacy system redesign reduced potential patient uses of OTC medications that were indicative of misuse (e.g., selecting an OTC that was contraindicated with existing health conditions or that differed from product labeling). That is, increased opportunities for pharmacy staff to engage with patients around medication safety issues, along with more visible cautionary signage and an OTC inventory comprising lower-risk medications when used to treat particular symptoms, generally decreased the occurrences of types of misuse to varying degrees. However, implementation of the intervention in more and different pharmacies, as well as assessing its impact on a greater number of patients, is warranted before the Senior Section can be considered a translatable and broadly valuable approach. As it now stands, the Senior Section represents a promising approach to enhance patients’ awareness of OTC risks and promote safe use of these medications through timesaving and effective pharmacy staff/patient interactions^28-29^ – a method that this research team is committed to continually evaluating and refining to achieve more universal application and sustained positive effect.

## Data Availability

No data are available.

## Acknowledgements

Study data were collected and managed using REDCap electronic data capture tools hosted at the University of Wisconsin-Madison, School of Medicine and Public Health (see Paul A. Harris, Robert Taylor, Robert Thielke, Jonathon Payne, Nathaniel Gonzalez, Jose G. Conde, Research electronic data capture (REDCap) – A metadata-driven methodology and workflow process for providing translational research informatics support, *J Biomed Inform*. 2009 Apr;42(2):377-81, for example). REDCap (Research Electronic Data Capture) is a secure, web-based application designed to support data capture for research studies, providing: 1) an intuitive interface for validated data entry; 2) audit trails for tracking data manipulation and export procedures; 3) automated export procedures for seamless data downloads to common statistical packages; and 4) procedures for importing data from external sources.

